# A Delay Differential Equation approach to model the COVID-19 pandemic

**DOI:** 10.1101/2021.09.01.21263002

**Authors:** I.N. Kiselev, I.R. Akberdin, F.A. Kolpakov

## Abstract

SEIR (Susceptible - Exposed - Infected - Recovered) approach is a classic modeling method that has frequently been applied to the study of infectious disease epidemiology. However, in the vast majority of SEIR models and models derived from them transitions from one population group to another are described using the mass-action law which assumes population homogeneity. That causes some methodological limitations or even drawbacks, particularly inability to reproduce observable dynamics of key characteristics of infection such as, for example, the incubation period and progression of the disease’s symptoms which require considering different time scales as well as probabilities of different disease trajectories. In this paper, we propose an alternative approach to simulate the epidemic dynamics that is based on a system of differential equations with time delays to precisely reproduce a duration of infectious processes (e.g. incubation period of the virus) and competing processes like transition from infected state to the hospitalization or recovery. The suggested modeling approach is fundamental and can be applied to the study of many infectious disease epidemiology. However, due to the urgency of the COVID-19 pandemic we have developed and calibrated the delay-based model of the epidemic in Germany and France using the BioUML platform. Additionally, the stringency index was used as a generalized characteristic of the non-pharmaceutical government interventions implemented in corresponding countries to contain the virus spread. The numerical analysis of the calibrated model demonstrates that adequate simulation of each new wave of the SARS-CoV-2 virus spread requires dynamic changes in the parameter values during the epidemic like reduction of the population adherence to non-pharmaceutical interventions or enhancement of the infectivity parameter caused by an emergence of novel virus strains with higher contagiousness than original one. Both models may be accessed and simulated at https://gitlab.sirius-web.org/covid-19/dde-epidemiology-model utilizing visual representation as well as Jupyter Notebook.

## Introduction

Mathematical modeling of the spread of infectious diseases is a powerful and widely used approach to predict infection, lethality and mortality rates in a certain country or over the world (Metcalf et al., 2020). It may also help reveal what should be the most effective administrative strategies and social containment measures in order to minimize loss of life and productivity and to curb the spread (Adam, 2020; Ciuriak and Fay, 2020; Cobey, 2020; Kucharski et al.,. 2020). The SIR (Susceptible, Infected and Recovered) method is an often used approach to build epidemiological models (Kermack and McKendrick, 1927). It has been widely applied to simulating the epidemic spread, control mechanisms and economic output of the COVID-19 in different countries (Atkeson, 2020; Calafiore et al., 2020; Fanelli and Piazza, 2020; Zhang et al., 2020). SIR-models can be easily extended, for example, to include different aspects of the disease. For instance, in (Banerjee et al., 2020) inclusion of the viral load and the impact on the immune human system into the SIR-model has enabled the identification of potential causes of two-phase exponential growth of the epidemic. Another natural extension is taking into account incubation time of the virus. Such models are usually called SEIR-models where E means exposed (Li and Muldowney, 1995).

SEIR-models found enormous application in theoretical studies of diverse aspects of the novel SARS-CoV-2 pandemic. In particular, such models were used to estimate the impact of different lockdown intensities on epidemic spread in China (Prem et al., 2020; Yang et al., 2020), United Kingdom (Ferguson et al., 2020) and Europe (e.g. the Netherlands) (Westerhoff and Kolodkin, 2020) for the year 2020, and even until 2025 for the USA, considering seasonal forcing and cross-immunity from the other betacoronaviruses (Kissler et al., 2020). The SEIR modeling has also been harnessed to estimate the effect of local and international travel restrictions on the spread of COVID-19 outbreak (Chinazzi et al., 2020).

The typical scenario of authorities’ actions (also called NPIs for Non-Pharmaceuticals Interventions) simulated in SEIR models is restriction on mobility and mass gatherings which reduce the number of contacts in the population and can be represented as an additional multiplier to the infection rate law reflecting social distancing (Westerhoff and Kolodkin, 2020). The control parameter may be changed by means of discrete events and piecewise functions. Other key NPIs are closing borders and quarantine on entry which diminishes the influx of infected individuals to the simulated region and mass testing for the virus where different modes of testing can be implemented in the model depending on the financial capabilities and government acts and policies (random tests or testing of infected with severe/critical symptoms, or considering contacts of infected one etc). A number of hospital beds and intensive care units (ICU) is another crucial factor in the fight of authority against COVID-19 which should be considered in epidemiological modeling (Tuomisto et al., 2020).

Despite the fact that initial results of the numerical study of SEIR models played an essential role in determining both basic laws of the primary development of the COVID-19 pandemic and core characteristics of the current pandemic situation, in the overwhelming majority this type of models uses mass action laws to describe the transitions between states (for example, from the incubation period to the symptomatic). Because of that, such models cannot always adequately reproduce the dynamics of such transitions. The methodological constraint of the SEIR models can be solved by using delayed differential equations which are able to explicitly capture the durations of the latent, quarantine, and recovery periods (Cooke and Van Den Driessche, 1996; Martcheva, 2015). Thus, Shayak and coathours numerically investigated the simplest retarded logistic equation with time delay to model the spread of COVID-19 in a city and demonstrated that solution of the model is significantly sensitive to small changes in the parameter values (Shayak et al., 2020). In the same time, more conventional SEIR-based delay differential equation models were proposed to reproduce the COVID-19 dynamics in Germany, China, South Korea, India and Japan (Götz and Heidrich, 2020; Menendez, 2020; Sharma et al., 2021; Utamura et al., 2020) and to predict the epidemic dynamics in Italy and Spain when it was in its early stages. However, these models did not take into account asymptomatic carriers and non-testing subpopulations as well as the progression of the disease’s severity.

Herein, we propose novel modification of the original SEIR model proposed in (Westerhoff and Kolodkin, 2020) using differential equations with weighted sums of delayed argument mixed with instant processes, which allow us not only model transition processes adequately to clinically observed data, but also directly quantify the proportion of hospitalized patients with moderate and severe symptoms, on an ICU, asymptomatic, tested and untested among them, which can be compared with the available statistics. The results of numerical analysis and model validation are demonstrated by the example of two European countries, Germany and France.

## Methods

### SEIR-like model

The overwhelming majority of SEIR-like models uses the mass action kinetic law for transitions between different stages of a disease(e.g. between exposed and infectious periods). Explicit drawbacks of this approach are that:

1. Parameters of those reactions are quite abstract and can not be easily related to real biological characteristics of the virus and
2. The model fails to correctly represent processes delayed in time. Here we will try to address those two problems.

In the current study we will use SBGN - Systems Biology Graphical Notation (Le Novere et al., 2009) for visual representation of mathematical models. Let’s consider a SEIR-like model with two levels of symptom severity.(Fig. 1).

**Fig. 1.**
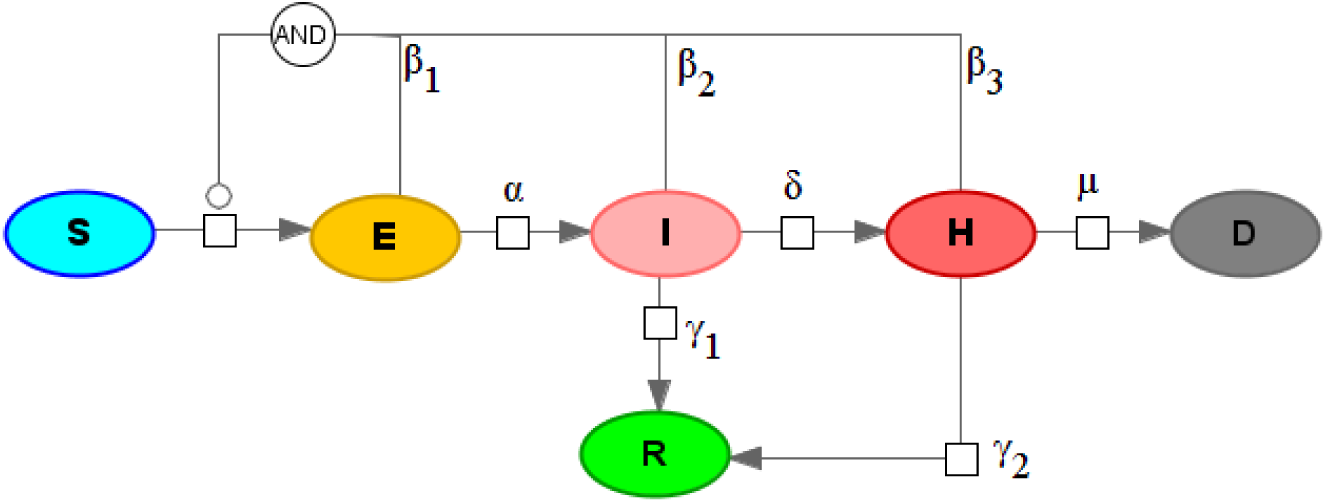
SEIRHD model with mass-action kinetics

Model equations:

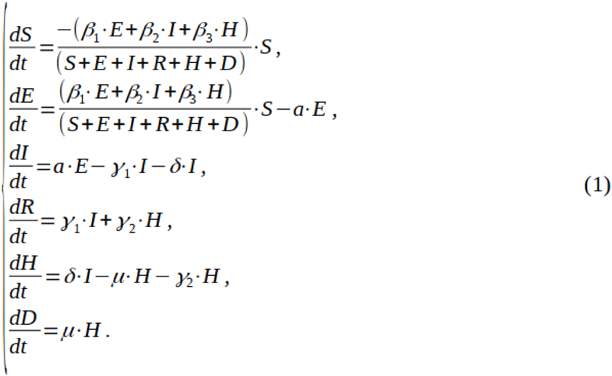

With initial values: *S*(0) = *S*_0_, *E*(0) = *E*_0_, *I*(0) = *R*(0) = *H*(0) = *D*(0) = 0, where S - susceptible population, E - exposed (in incubation period) population, I - infected (with mild symptoms) population, H - infected (with severe symptoms), R - recovered population, β_1_, β_2_, β_3_ - infection rates for different contagious groups, a - symptom onset rate, δ-symptoms worsening rate, µ - death rate, γ_1_, γ_2_-recovery rate (for mild and severe symptoms respectively).

### Duration of processes

A numerical value of the parameter *a* in the model (1) is related to the median incubation period in the population. For example if we set *a* = ln(2)/5.1 then 50% of individuals who were exposed to the virus at time t = 0 will become symptomatic at time t = 5.1 days.

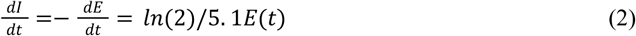

Distribution of incubation period in this model compared to the experimental data from (Lauer et al., 2020) is presented in Figure 2. A One can easily observe that it is inconsistent with statistical data on incubation period. For example, for almost 10% of the infected, symptoms will onset in one day after exposure.

**Fig. 2.**
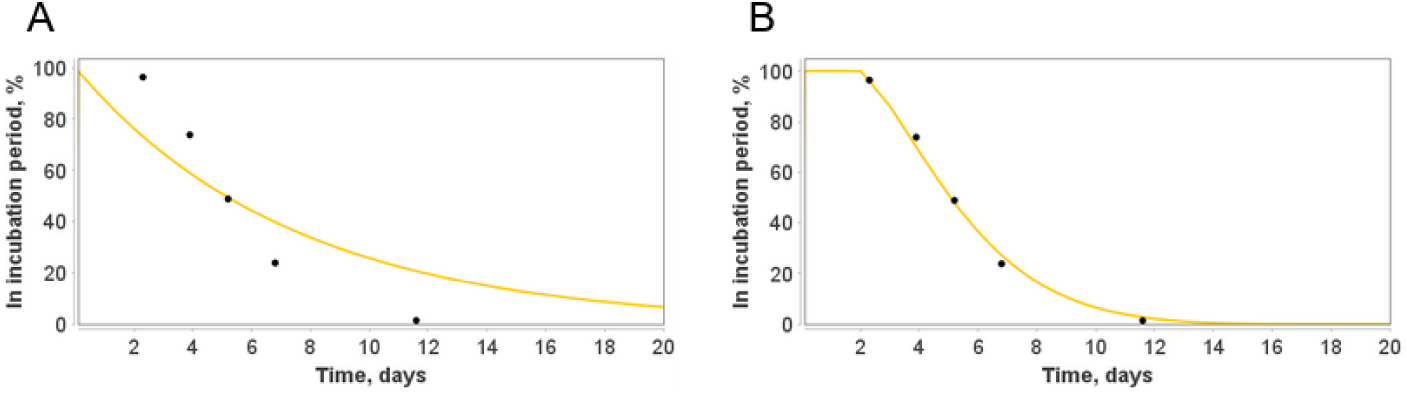
A comparison between two models of the incubation period: (A): based on the mass-action kinetics law (2). B: using weighted sum of delays (4). Statistical data of the incubation period quantiles are taken from (Lauer et al., 2020).

Unfortunately, having only one parameter *a* we can not fit the curve to this statistical data. This is also the case for other processes delayed in time with certain distribution of length. A possible solution to overcome the issue is use of different forms of the kinetic law:

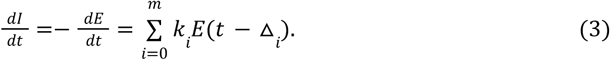

Herein, we use a weighted sum of the delayed number of exposed individuals. We have a 2*m parameters which can be estimated to reproduce an experimental data. Typically, m = 1 or m = 2 is enough to comprehensively fit the data, keeping the number of parameters reasonably low.

For example, to fit the data from (Lauer et al., 2020) we employ two delays only:

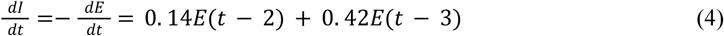

Simulation results of the incubation period’s model demonstrating differences between theoretical curves obtained using two methodologies are presented in Figure 2.

Another benefit of the time-delay based approach is an opportunity to reproduce an experimental data on diverse epidemiological processes (incubation period, recovery, worsening of symptoms from mild to severe and others) once and separately from the rest of the model structure based on the known distributions of duration of those processes for particular infectious disease only.

### Competing processes

Another opportunity to improve the original model and bring it closer to reality is to consider different possible transitions from the same subgroup. For example, infected patients may either recover or progress to severe symptoms (Fig. 1). In that case parameters δ and γ_1_ are related not only to durations of corresponding processes but also with recovery rate for mild symptomatic (or fraction of severe symptomatic among mild symptomatic). An issue with two alternatives arises when the fast process has less probability and therefore less fraction of patients involved in this direction of the infectious process. According to the statistics (Boelle et al., 2020), the process of worsening of symptoms (median time is 5 days) is faster than recovery (median time equals to 14 days) implying that δvalue should be larger than γ_1_ value. As in the previous subsection, we may set these parameters as δ = *ln*(2)/5, γ_1_ = *ln*(2)/14. However, in that case (Fig. 3) the model will show that the fraction of patients that transit to the severe symptoms is larger than the fraction recovered which is not the case in reality (WHO report, 2020).

**Fig. 3.**
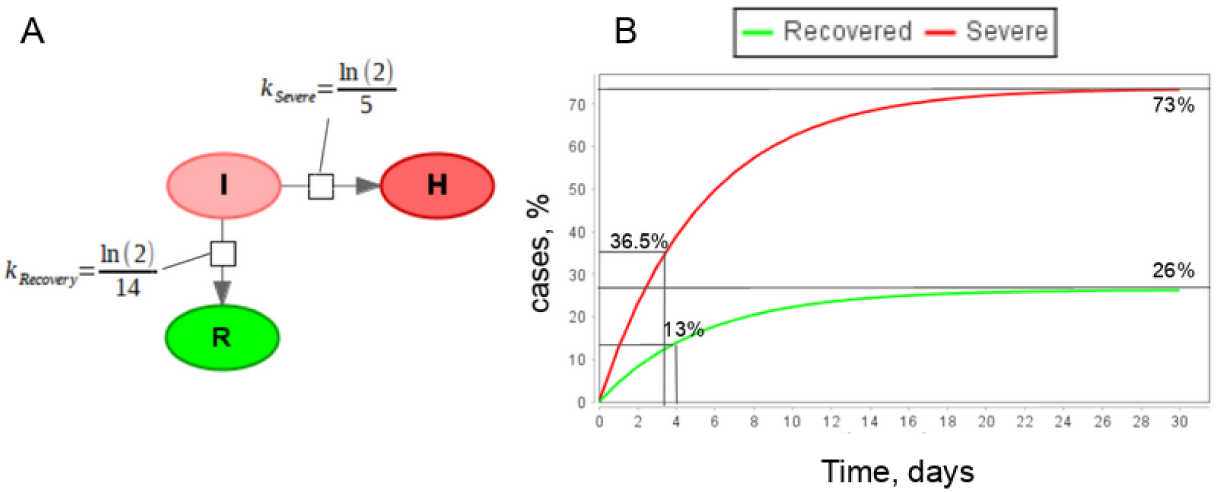
The simple mass-action model with two competing processes. (A): SBGN representation of the model, here I - infected individuals, R - recovered, H - patients with severe symptoms; (B): Simulation results of the model.

The possible solution to overcome the discrepancy is to consider those two processes separately. Firstly, patients will instantaneously transit from “Symptomatic” to “Symptomatic who will recover in future” (*I*_*R*_) and then from *I* to R. In similar way another fraction of symptomatic patient will instantaneously transit to “Symptomatic who will need hospitalization” (*I*_*H*_) and only afterwards from *I*_*H*_ to *H*. The updated model is presented in Figure 4 and corresponding model equations are:

**Fig. 4.**
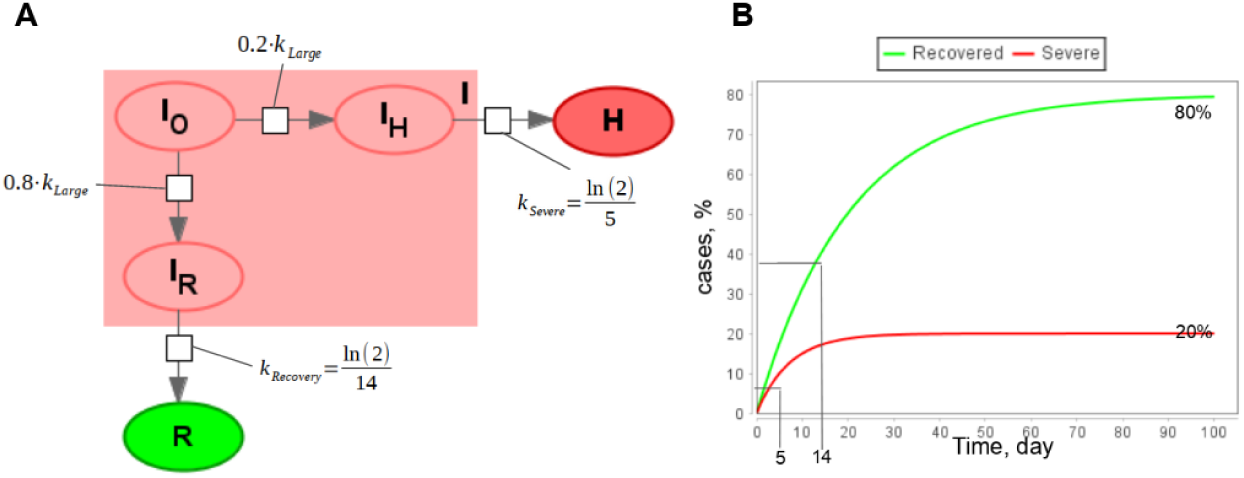
A part of the epidemiological model with alternative competing processes. (A): Visual representation in SBGN format; (B): Simulation results of the model for fractions of recovered and patients with severe symptoms.

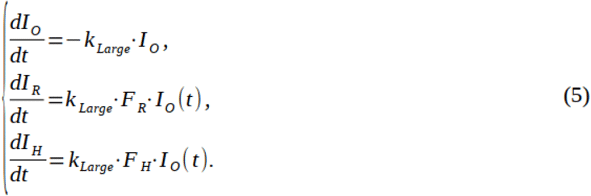

Where *F*_*R*_ = 0. 2 - a fraction of infectious individuals who will not have worse symptoms.

*F*_*H*_ = 0. 8 - a fraction of infectious individuals who will have worse symptoms.

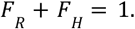

*K*_*Large*_ - constant value which is large enough to render reactions instant.

The total number of individuals with mild symptoms is calculated as a sum of all subgroups:

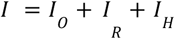

This technique can be combined with delayed equations described in the previous section. Thus, we can construct a version of the model (1) taking into account the duration of some infectious processes and the existence of competing processes. The final version of the model is presented in Figure 5.

**Fig. 5.**
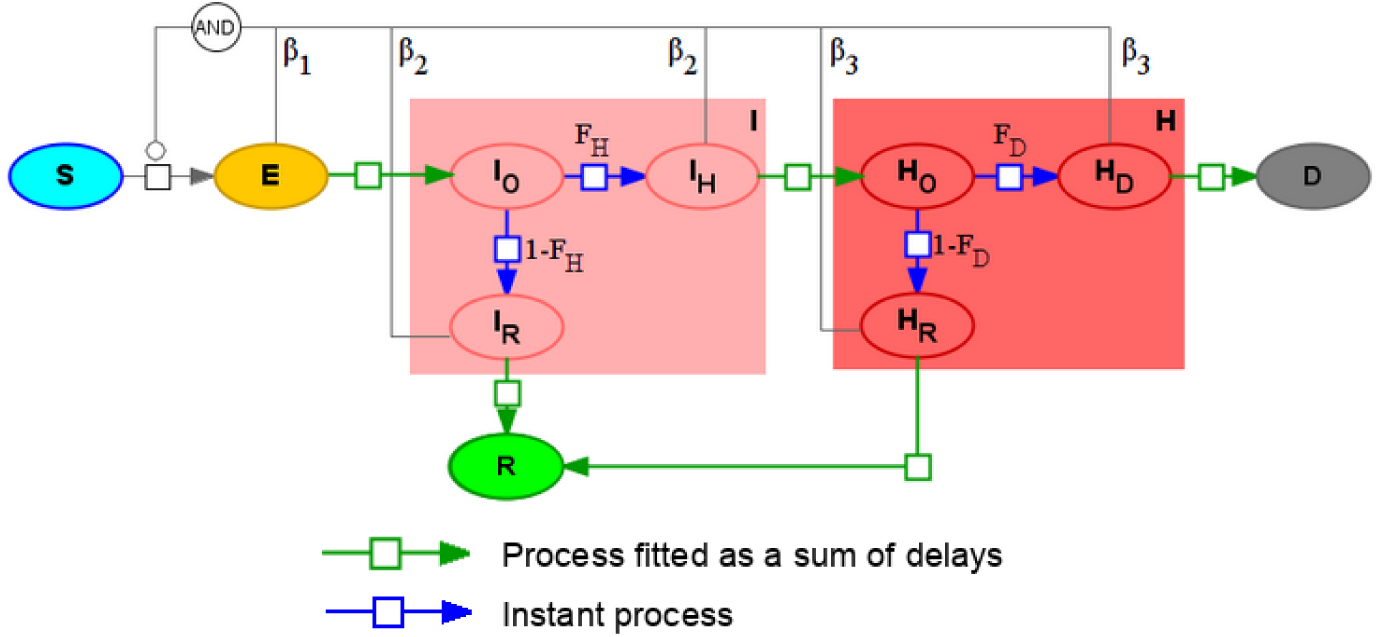
SEIR-like model with instant and delay-based processes (2)

Advantage of the updated model is that instead of 8 parameters which do not explicitly correspond to real characteristics of infectious processes and has to be fitted to experimental data we have only three parameters, two fraction parameters: *F*_*H*_ - fraction of symptomatic individuals with severe symptoms, *F*_*D*_ - disease lethality that can be drawn from statistical data and processes that are fitted to experimental data separately from the rest of the model. Comparison is given in Table 1.

**Table 1.** Comparison of parameters in models (1) and (2)

| Modeled process | Parameters |  |
| --- | --- | --- |
|  | SEIRHD Model (1) | Delay-based Model (2) |
| Exposure to the virus | Infection rates $\beta_1, \beta_2, \beta_3$ | Infection rates $\beta_1, \beta_2, \beta_3$. |
| Incubation period | $\alpha$ | Fitted process. |
| Recovery with mild symptoms | $\gamma_1$ | Fraction of severe symptomatic $F_H$, Fitted process. |
| Progress to severe symptoms | $\delta$ | Fraction of severe symptomatic $F_H$, Fitted process. |
| Recovery with severe symptoms | $\gamma_2$ | Disease lethality $F_D$, Fitted process. |
| Death | $\mu$ | Disease lethality $F_D$, Fitted process. |

### Initial model

As a basis for our model we used the previously created SEIR model (Westerhoff and Kolodkin, 2020) of the Covid-19 epidemic. This model differs from the most SEIR models by differentiating between tested and non-tested infected subjects. It was created in the Systems Biology software COPASI (Hoops et al., 2006) which allows one to specify the kinetics of the process mechanistically. COPASI translates these specifications into differential equations which it integrates either as a function of time, or by requiring steady state. The software honors restrictions as specified in terms of algebraic equations and ‘events’ which instantaneously change numeric values of the model parameters triggered by logical expressions transiting from “false” to “true”. COPASI models are SBML compatible, and can be exported into the format. The latter greatly facilitates model reuse and reproduction.

### Data sources

Statistical data for Germany and France was taken from Our World In Data web site (https://ourworldindata.org/). This web-portal provides the data on the total number of cases, new cases each day, number of hospitalized, transferred to ICU patients, vaccinated individuals and total number of deaths.

To take into account statistical data on government actions in the model we employed the Stringency Index developed by the Blavatnik School of Government of the University of Oxford (Hale et al., 2021) which incorporates data on 10 different measures corresponded to: C1 - School closing, C2 - Workplace closing, C3 - Public events cancellation, C4 - Restriction on gatherings, C5 - Public transport closing, C6 - Stay at home requirements, C7 - Internal movement, C8 - International movement, H1 - public information campaigns.

### BioUML Platform

BioUML (http://www.biouml.org) used in the study is an integrated Java platform for modeling of biological systems (Kolpakov, 2019). It supports different mathematical formulations for the model development including ODE, delay-based, algebraic systems, discrete events, agent-based, and stochastic modeling. The platform also incorporates a module for the automatic and manual parameter fitting to an experimental data. Models developed in the BioUML are based on main standards in systems biology: 1) SBML - Systems Biology markup Language (Hucka et al., 2018) for mathematical description and 2) SBGN for visual representation. A model can be built and edited in the platform as a visual diagram (e.g. in SBGN notation) based on which a Java code is generated for model simulations. Additionally, BioUML is integrated with Jupyter hub (https://jupyter.org/) for interactive data and model analysis as well as an essential and user-friendly tool for reproducibility of the simulation results.

## Results

### Model structure

The final version of the proposed delay differential equations (DDE) model consists of the next set of subpopulations or groups (Fig. 7):

**Fig. 6.**
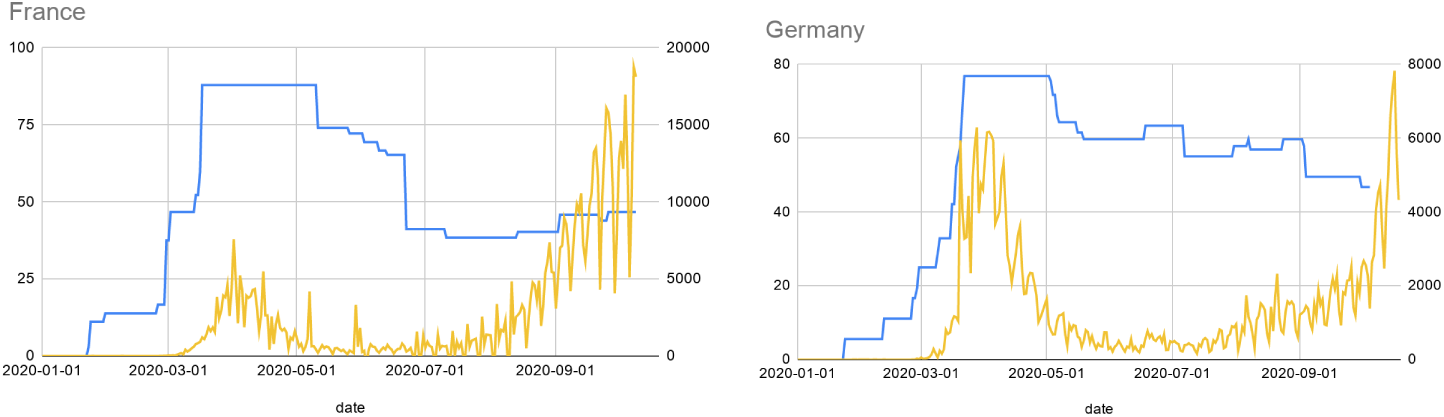
Stringency of government measures (blue) and new reported cases per day (yellow)

**Fig. 7.**
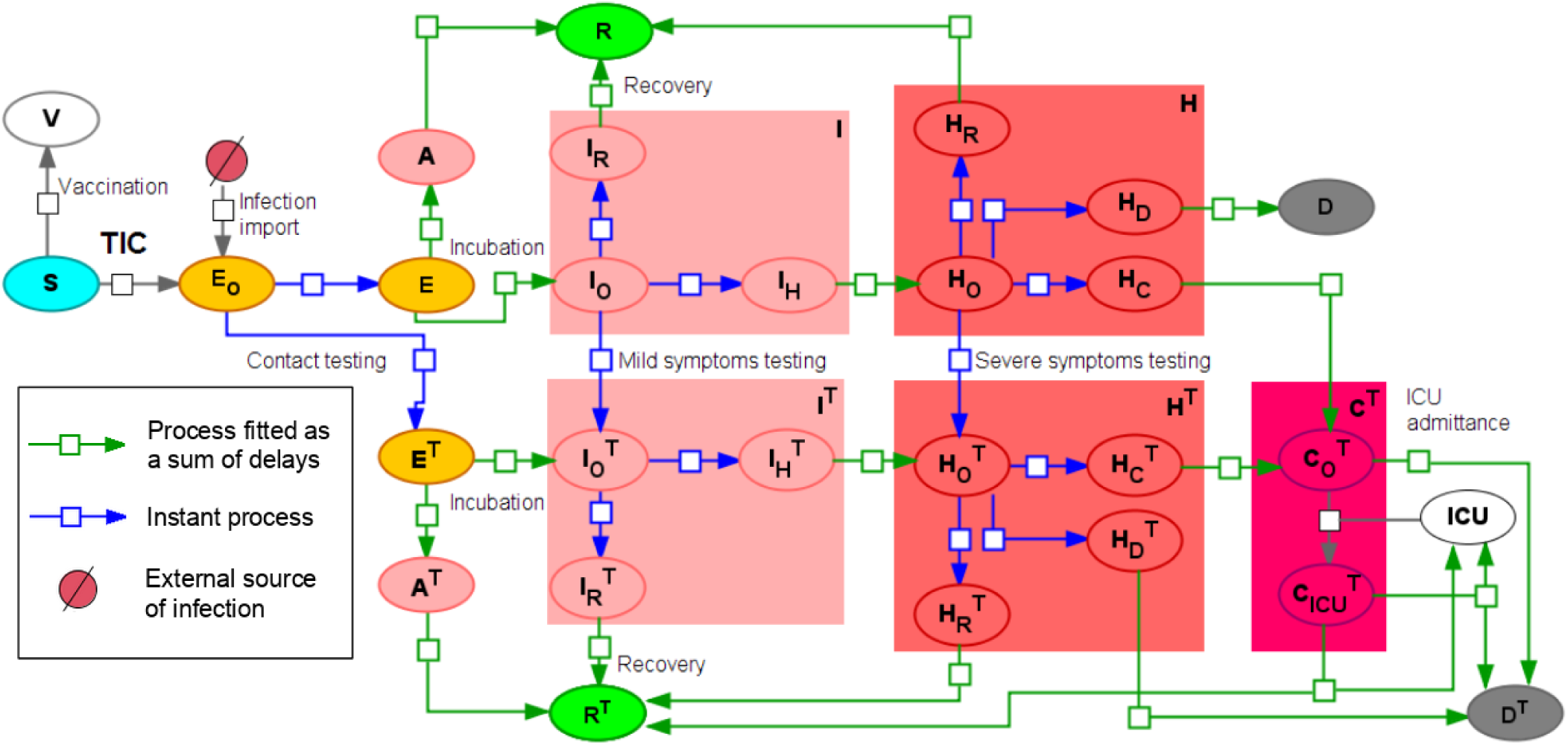
Overall SEIR-like model with instant and delayed processes.

1. *S*- Susceptible to the SARS-CoV-2 virus.
2. *N* - Non susceptible due to previously existing immunity (Doshi, 2020; Mateus et al., 2020; Ng et al., 2020; Pinto et al., 2020; Shrock et al., 2020). Those individuals can not be infected and can not infect anyone else accordingly. Note that this group does not include vaccinated individuals.
3. *V*- vaccinated subpopulation, considered to be immune to the virus.
4. *E* - exposed to the virus. After the incubation period they will transit either to asymptomatic or symptomatic. Here we do not use additional subgroups due to equal time intervals for both transitions.
5. A - asymptomatic individuals, which will recover over time but can infect others (Oran et al., 2021).
6. *I* - mild symptomatic group. It comprises three subgroups: with onset symptoms (*I*_*O*_) then they are instantly divided into those who will recover (*I*_*R*_) and those who will progress to the severe symptomatic (*I*_*H*_). Transition is done according to the fraction of severe symptomatic among those who show any symptoms (*F*_*H*_).
7. *H* - severe symptomatic group which comprises four subgroups: 1) with just onset symptoms (*H*_*O*_). They instantaneously transit into subgroups of individuals who will eventually recover (*H*_*R*_), die (*H*_*D*_) or progress to critically ill (*H*_*C*_). Transitions are performed according to the disease lethality (*F*_*D*_) and the fraction of critically ill (*F*_*C*_).
8. *C* - critically ill group where ICU is required in order to recover. If no ICU is available these patients will die. All critically ill patients are considered to be automatically tested for the virus infection.
9. *R* - recovered.
10. *D* - dead.
11. All infected subgroups (except critically) also have registered or tested counterparts: *A*^*T*^, *E*^*T*^, *I*^*T*^, *H*^*T*^, *R*^*T*^, *D*^*T*^. In the model patients may be tested at three different stages:
  1. When being exposed to the virus. It is done through contact tracing procedures. Percent of exposed to the virus who will be tested and registered is set by *T*_*E*_ parameter.
  2. Upon symptoms onset. Percent of mildly symptomatic individuals who will be registered is given by parameter *T*_*I*_.
  3. Upon severe symptoms onset. Percent of severely symptomatic individuals who will be registered is given by parameter *T*_*H*_.

Most transitions in the model are described as either instant processes or as preliminary fitted processes (blue and green arrows, correspondingly, in Fig. 7). To fit delayed processes we used data from (Lauer et al., 2020) for incubation period and (Boelle et al., 2020) for other processes. We also harnessed data provided by Our World in Data for France for recovery\dying in hospitals. To this end we constructed a partial model describing the process of admitting hospital, transition to ICU and leaving hospital (Fig. 8). As one can see in Table 2, obtained quartiles are quite different from those presented in (Lauer et al., 2020). Givendaily numbers of hospital admission, daily number of ICU admissions and daily number of hospital patients we fitted the process of leaving hospital utilizing formula (3). It should be noted that we assume that all severely ill patients are tested and moved to the hospital (*T*_*H*_ = 1). However, it is not always the case and should be addressed in the updated version of the model. Overall scheme of the partial model in SBGN format as well as a result of the model fitting to data on hospitalization in France are presented in Fig. 8.

**Table 2.** Processes fitted using formula (3) and data from (Boelle et al., 2020; Lauer et al., 2020). More details are provided in supplementary materials (Supplementary figs. 1-7).

| Process | Median [Quartiles] (Observed) | Calculated quartiles | Results of the fitting |
| --- | --- | --- | --- |
| Incubation period | 5.1 [2.2, 3.8, 5.1, 6.7, 11.5] (Lauer et al., 2020) | [2.13, 3.56, 5, 6.9, 11.6] | $0.14 \cdot x(t - 2) + 0.04 \cdot x(t - 3)$ |
| Recovery with mild or no symptoms | 14 [8, 20] (Boelle et al., 2020) | 14.9 [8.13, 21.2] | $0.048 \cdot x(t - 3) + 0.0017 \cdot x(t - 4) + 0.016$ |
| Onset of severe symptoms | 5 [2, 9] (Boelle et al., 2020) | 4.9 [2, 9] | $0.14 \cdot x(t) + 0.014 \cdot x(t - 5)$ |
| Release from hospital (without ICU) | 9 [6, 15] (Boelle et al., 2020) | 9.2 [5.8, 14.7] | $0.086 \cdot x(t - 3) + 0.67 \cdot x(t - 14.9)$ |
| | 13.8 [5.7, 27.7] Fitted based on hospitalization data from ourworldindata.org | 13.8 [5.75, 27.7] | $0.051 \cdot x(t)$ |
| Onset of critical symptoms | 1 [0, 2] (Boelle et al., 2020) | 0.96 [0.4, 1.9] | $0.717 \cdot x(t)$ |
| Discharge from ICU | 20 [10, 39] (Boelle et al., 2020) | 20.4 [9.6, 38.7] | $0.007 \cdot x(t - 3.583) + 0.028 \cdot x(t - 1.585)$ |

**Fig. 8.**
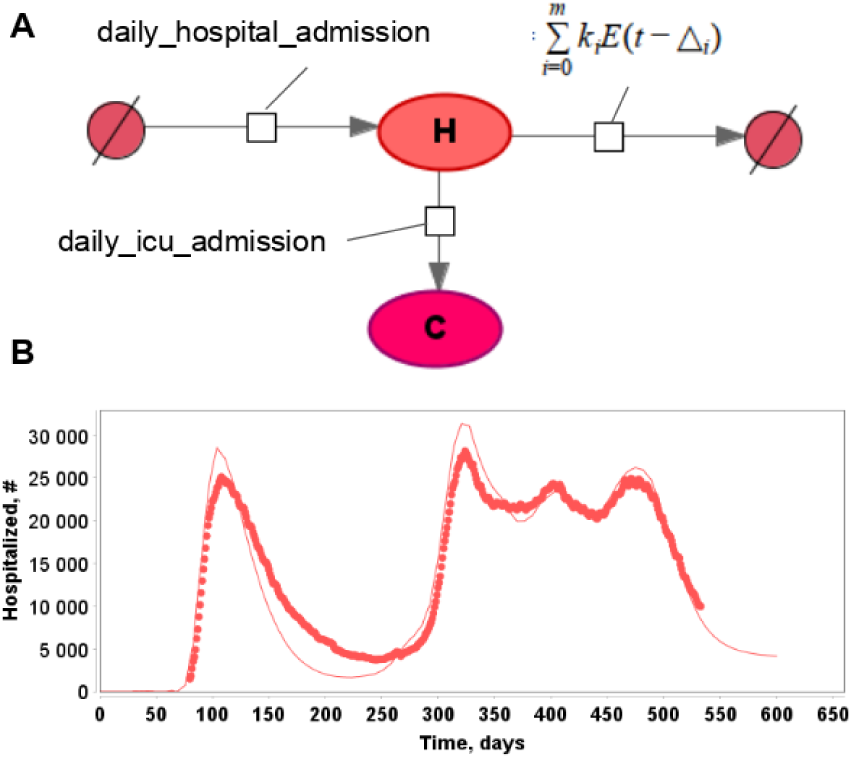
(A): Partial model of the hospitalization due to Covid-19 using delay equations for hospital release. Data on daily hospital and ICU admission taken from ourworldindata.org. (B): Results of the model fitting to the number of hospitalized patients not in ICU in France.

There are also three transitions in the model treated differently:

1. **Vaccination**. Based on the known statistical data we established the number of individuals vaccinated each day in a certain country. This number is divided proportionally between all subgroups eligible for vaccination which are susceptible, non-susceptible and recovered (registered and not registered) individuals. Among them, vaccination of susceptible subpopulation only affects the model dynamics and thus is presented on the diagram and model equations. The kinetic law for this process is zeroth-order: *dV*/*dt* = *k*_*V*_ with *k*_*V*_ changed each day based on the tabular data for a country. We also assume that vaccines have 100% efficiency, and the vaccinated can not be infected in the current version of the model.
2. **ICU admittance**. We considered this process to require a free ICU and be instant in most cases. However, lower value of the kinetic constant may be used to reflect the fact that not everyone who needs ICU gets it:

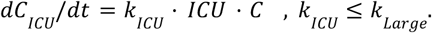
3. **Process of infection**. Transition from susceptible to exposed is defined using Total Infection Coefficient (*TIC*) which we calculated according to the original version of the SEIR model (Westerhoff and Kolodkin, 2020):

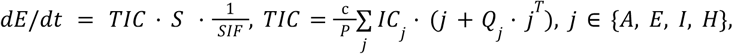

where *c* - average number of contacts for individuals per day in the simulated region which can be drawn from statistical surveys, *IC*_*j*_ - a probability to be infected by a member of the corresponding group upon contact, *Q*_*j*_ - quarantine coefficient for corresponding group. Only registered individuals are subject to quarantine, *SIF* is a stringency index factor calculated based on the Stringency Index which reflects government NPIs and imposed limits such as mask regime, limit on mass gatherings, school closing etc. Stringency index ranges from 0 (no interventions) to 100 (maximum possible interventions). In the current study we recalculated it to SIF as follows:

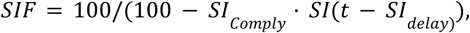

Where *SI*_*Compy*_ ∈ [0, 1] identifies extent of the population compliance to government interventions, *SI*_*delay*_ - time delay between government interventions enacting and their effect.

### Simulation results

The final version of the DDE based model (Fig. 7) was fitted to Covid-19 epidemic data in Germany and France from 01.01.2020 (model time t = 0) to 23.08.2021 (model time t = 600). We have divided the overall time duration into three intervals: first, second and third pandemic waves. Values of some model parameters were changed between waves to reflect changes in the properties of the disease and reaction to it. All model parameters whose values were different between models fitted to the data on two countries or varied during the pandemic are presented in Table 4. Simulation results with corresponding statistical data for Germany and France are presented in Figures 9 and 10.

**Table 3.** Model parameters. * means value depends on the model region (see Table 4).

| # | Symbol | Description | Value | Units | Source |
| --- | --- | --- | --- | --- | --- |
| 1 | $S_F$ | Fraction of susceptible to the virus in the population | * | % | Statistics |
| 2 | $A_F$ | Fraction of asymptomatic among susceptible | 50 | % | Statistics |
| 3 | $H_F$ | Fraction of severe among mild | * | % | Statistics |
| 4 | $C_F$ | Fraction of critically ill among severe | * | % | Statistics |
| 5 | $D_H$ | Fatality rate for severe symptoms | * | % | Statistics |
| 6 | $D_{ICU}$ | Fatality rate for patients in ICU | * | % | Statistics |
| 7 | $T_E$ | Percent of registered while in incubation period or asymptomatic | * | % | Fitting |
| 8 | $T_M$ | Percent of registered while having mild symptoms | * | % | Fitting |
| 9 | $T_H$ | Percent of registered with severe symptoms | 100 | % | Fitting |
| 10 | $Q_E$ | Mobility limit for registered in incubation period | 0.7 | - | Fitting |
| 11 | $Q_A$ | Mobility limit for registered asymptomatic | 0.7 | - | Fitting |
| 12 | $Q_H$ | Mobility limit for registered with severe symptoms | 0.1 | - | Fitting |
| 13 | $Q_M$ | Mobility limit for registered with mild symptoms | 0.27 | - | Fitting |
| 14 | $IC_E$ | Probability of being infected upon contact with exposed in incubation period | 5.6 | % | Fitting |
| 15 | $IC_A$ | Probability of being infected upon contact with asymptomatic | 3.4 | % | Fitting |
| 16 | $IC_M$ | Probability of being infected upon contact with mild symptoms | 3.1 | % | Fitting |
| 17 | $IC_H$ | Probability of being infected upon contact with severe symptoms | 1.8 | % | Fitting |
| 18 | $CF$ | Average number of contacts per day for one individual | * | # | Statistics |
| 20 | $SI_{Delay}$ | Delay between government interventions enacting an their effect | 3.8 | day | Fitting |
| 21 | $SI_{Comply}$ | An extent of the population compliance to government interventions | * | - | Fitting |

**Table 4.**
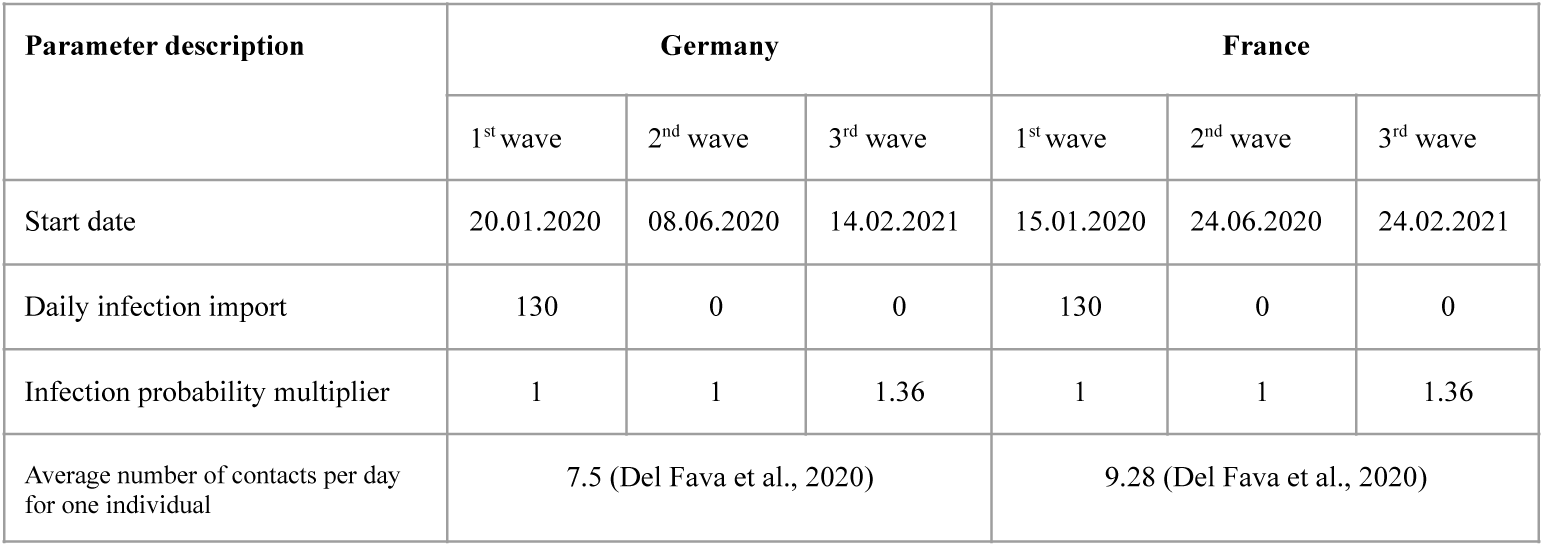
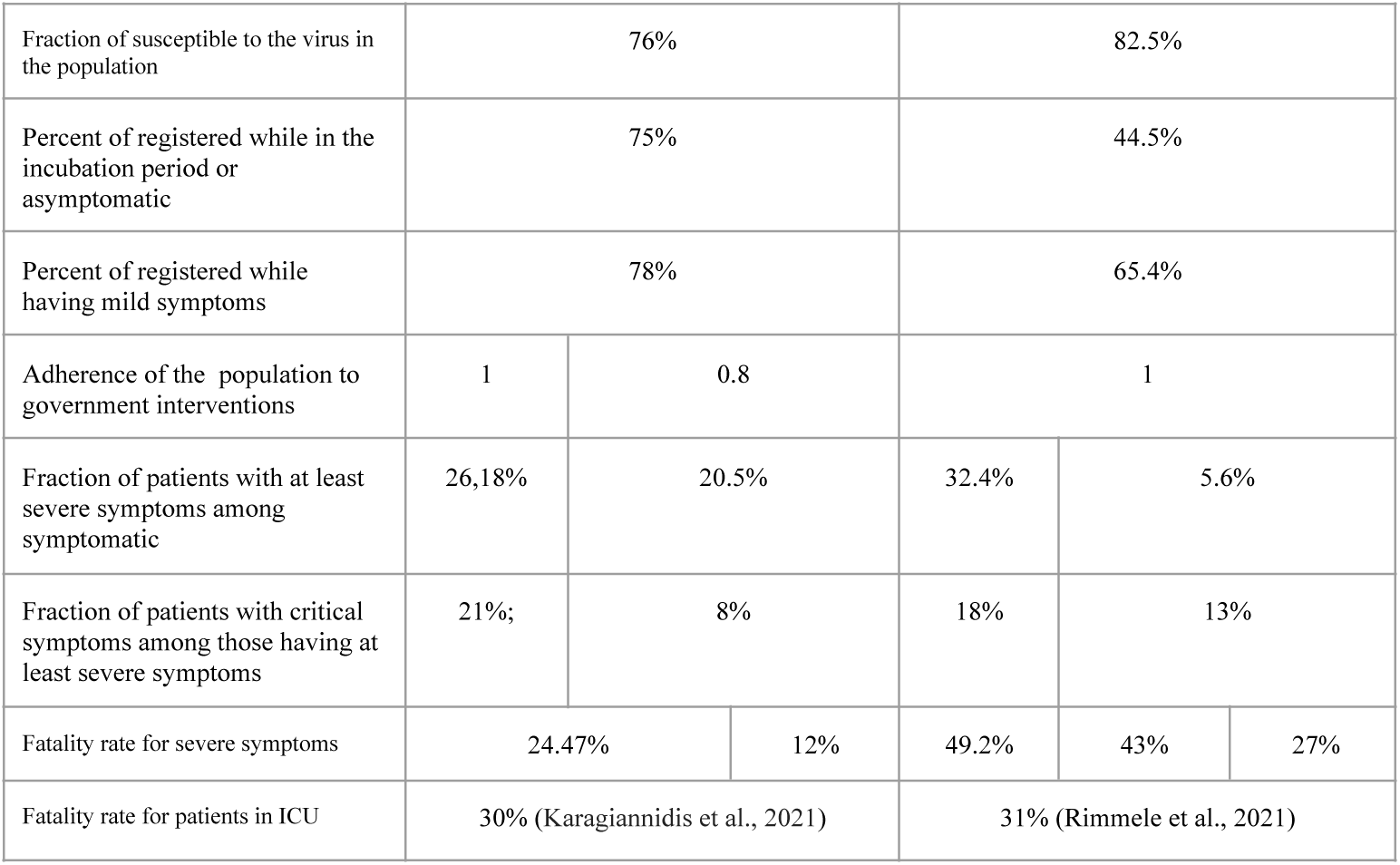
Estimated parameter values.

**Fig. 9.**
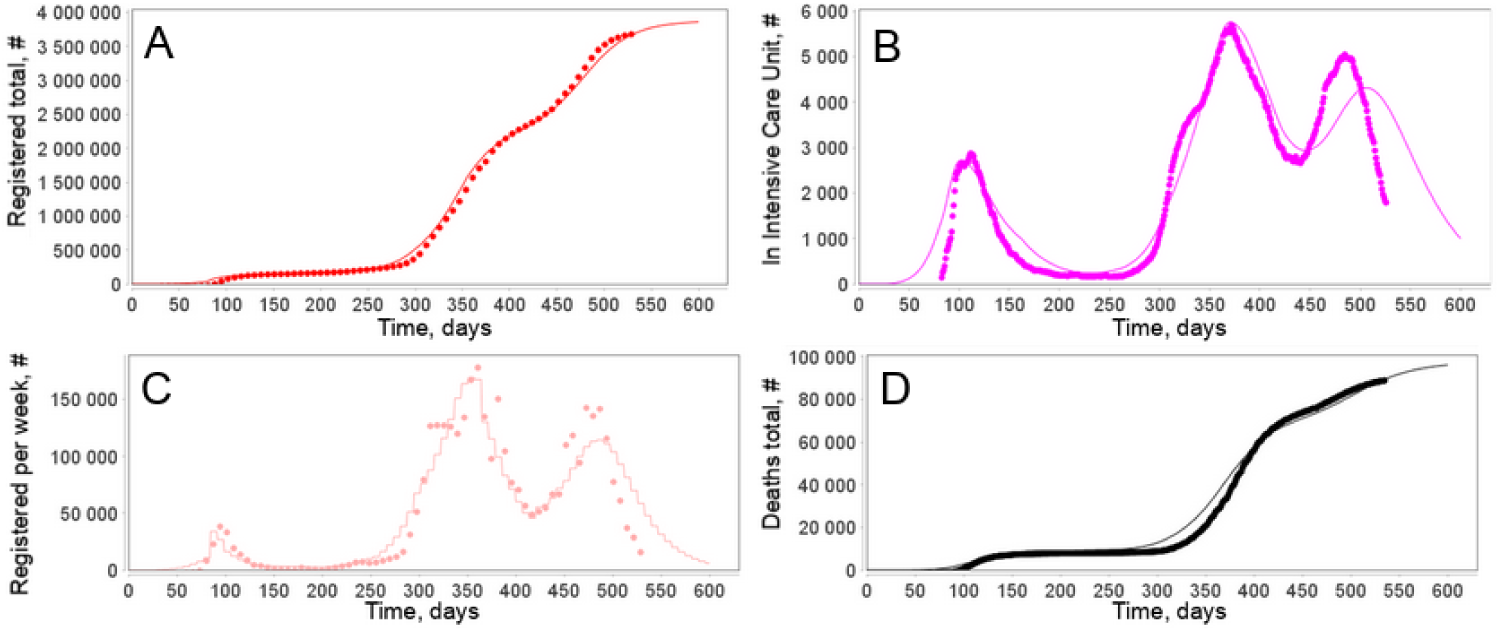
Simulation results and corresponding statistics for Germany. (A): Registered in total. (B): In ICU. (C): Registered per week. (D): Deaths due to Covid-19 in total. Time is counted from the start of 2020, t = 1 means 01.01.2020. Statistics were taken from ourworldindata.org web-site.

**Fig. 10.**
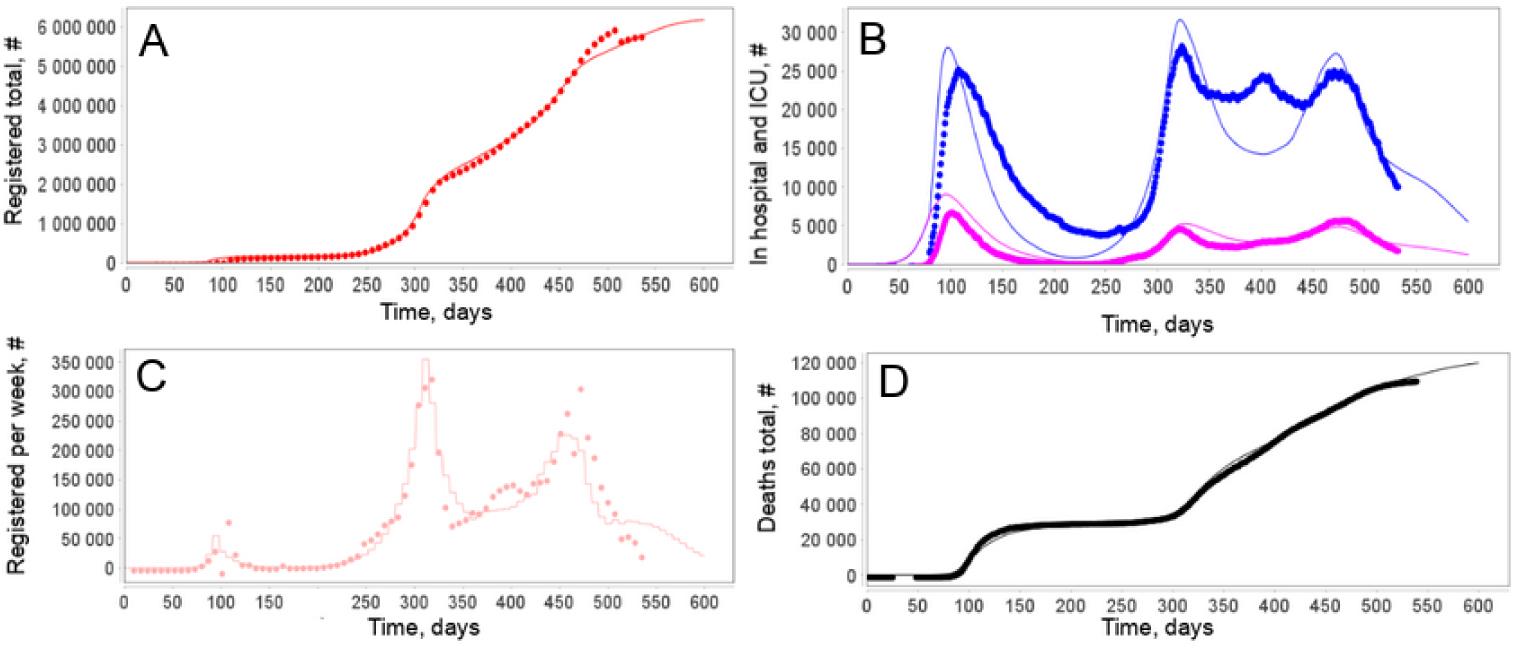
Simulation results and corresponding statistics for France. (A): Registered in total. (B): The number of hospitalized patients (blue) and in ICU (pink). (C): Registered per week. (D): Deaths due to Covid-19 in total. Time is counted from the start of 2020, i.e. model time point t = 1 means 01.01.2020. Statistics were taken from ourworldindata.org web site. It should be noted that 346261 cases were officially deducted from the total number of registered cases on 20 May 2021 because they were counted twice.

**Fig. 11.**
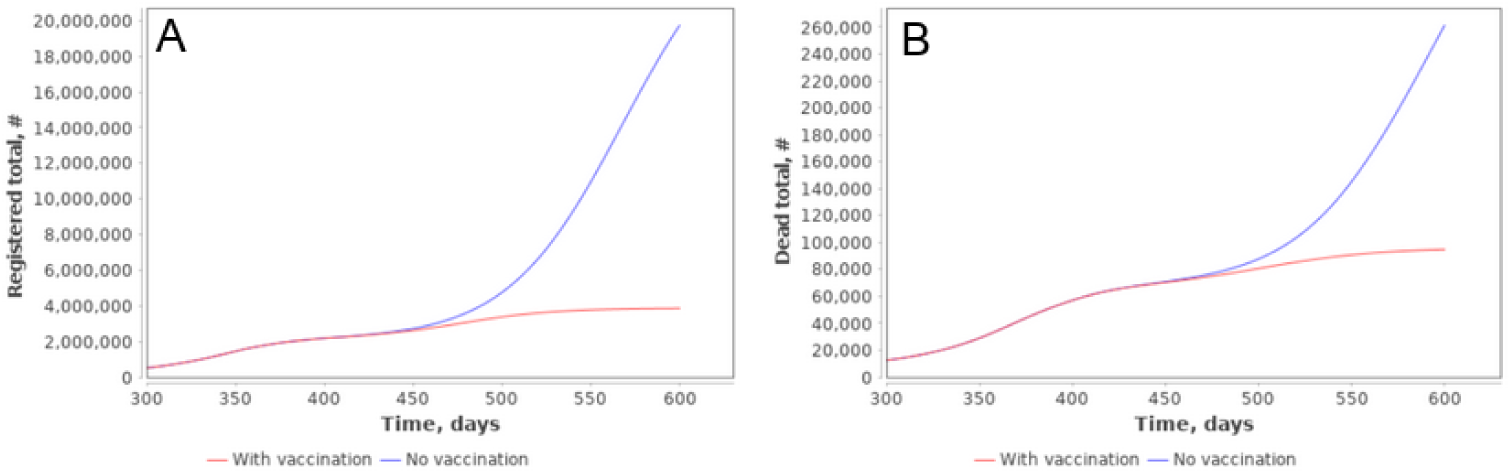
Simulation results for Covid-19 epidemic in Germany with current level of vaccination (red) and without vaccination (blue). (A) Registered in total. (B) Deaths due to Covid-19 in total.

1. The First wave: This interval starts somewhere in January 2020. From this time point infected individuals started to arrive in the country in significant amounts. Patient zero in Germany entered the country on 20 January and was registered on 27 January (Bohmer et al., 2020). We assumed in the model that import of infection to Germany has begun since 20 January. The import rate was estimated to be 130 infected individuals per day. This import was ended on 16 March 2020 (t = 76), when the European Union as a whole announced the closure of all its external borders to non-citizens (European Commission, 2020). For France the first case was identified on 24th January. However, individuals infected by SARS-CoV-2 were present as early as December 2019 according to some sources (Deslandes et al., 2021; Carrat et al., 2021). Unfortunately, we do not have data on how many infected individuals arrived in France or Germany before borders were closed on 16 march 2020. Thus, for any starting date we may obtain the number of infected arriving per day to fit the same observable data. We kept the same value of 130 persons per day for France and estimated the start of mass influx to France on 15 January. In the current study we have added a constant influx of exposed individuals (E) in the model with constant rate *Import*_*Initial*_. Of course, this process was more complicated in reality. Eventually, the number of new cases falls in both countries in the first half of the year.
2. Second wave - starting from summer 2020 the number of new cases began to rise again implying the second epidemic wave in the region with many more registered cases. It may be attributed to relaxing anti epidemic restrictions which can be traced by lower level of Stringency Index. In the France model it caused a second wave in accordance with existing statistics. For Germany, however, the model fitted for the first wave was not able to predict the second wave. Thus, Stringency Index in the form currently used in the model can not accurately predict the second wave in Germany. Possible solution was to decrease population compliance to government interventions *SI*_*Comply*_ from 1 to 0.8. Despite a larger number of cases - number of deaths and ICU admissions did not rise as significantly. According to (Karagiannidis et al., 2021) the number of patients requiring ICU dropped by 50% during the second wave. This may be attributed to changing in patients’ average age or better treatment procedures. In both models it was reflected by changing next parameters: the fraction of severely ill among symptomatic as well as the fraction of critically ill among severely ill and mortality of patients with severe symptoms.
3. Third wave - starting from February 2021 a new significant rise in numbers of cases begins in both countries. It may be connected with the spread of new strains of the virus. Indeed, new virus lineage with additional mutations in the spike region, B.1.1.7 strain (Frampton et al., 2021), was rapidly spreading in some European countries at this time period (Hodcroft, 2021). New strains are much more contagious but less deadlier (Andersson et al., 2021; Kirby, 2021; Sheikh et al., 2021). However, the latter is debatable and discussed below. This was modeled by multiplying all probabilities to be infected upon contact by the same multiplier. The multiplier’s value was fitted for both countries separately, but the obtained numerical value was the same - probability increased by 36%.

It should be noted that although there are estimates of the proportions of different severity of symptoms of the disease, nevertheless, these probabilities strongly depend on the patient’s age and, therefore, for each individual country, these proportions may differ. In addition, according to statistics, these parameters have changed over time (possible reasons: a change in the age composition of patients, the development of new therapies in hospitals, mutations and the emergence of new strains of the virus). For these reasons, in the current study, these parameters were the objects of assessment on a country-by-country basis.

As can be seen from simulation results (Figures 9 and 10) the model accurately reproduces the reported new cases per week and total number of cases as well as the number of hospitalized patients on ICU and total deaths in each country over time of the pandemic.

To assess the impact of vaccination to control COVID-19 burden in Germany we applied the proposed DDE fitted model. Number of people vaccinated each day was brought from ourworldindata statistics, according to which Mass vaccination in Germany began 27 December 2020 which corresponds to model time t = 363 (while t = 1 corresponds to 1 January 2020. It should be noted that we consider every individual administered with at least one dose of the vaccine as completely immune to the virus. According to the model predictions, the vaccination campaign and coverage in Germany significantly reduces the cumulative numbers of cases and deaths, while a no-vaccination scenario would lead to approximately 5 times more infected cases and 2.5 times more deaths in this country

## Discussion

We have proposed the methodology to overcome some shortcomings of the classic SEIR-based epidemiological models via the novel epidemiological model which utilizes DDEs to take into account different time scales of epidemiological processes and instantaneous splitting procedure to describe competing processes. The essential benefits of the developed model comprises:

1. Most of the epidemiological processes (symptoms onset, recovery, dying etc.) are described using kinetic laws with delayed arguments. These modeling processes can be fitted separately from the rest of the model and applied kinetic laws provide more precise reproduction of real properties of those processes than mass-action laws with a single parameter.
2. If a model has two or more competing transitions, a division into separate subpopulations removes their undesired mutual influence and allows to simulate fast and slow transitions with correct fractions of patients undergoing each transition.
3. Model parameters that are not parts of fitted processes described previously have direct mechanistic meaning (i.e. disease lethality, susceptibility, probability of different symptoms severity) and can be drawn from the statistics.

Combination of these advantages makes the model more reliable with real properties of the pandemic and eliminates most of the abstract parameters usually used in SEIR-like models. It is worth to note there are other studies addressing these issues using Erlang distribution (Arino and Portet, 2020) and delayed equations (Devipriya et al., 2021; Yang et al., 2021). However, to our best knowledge, there are no publications demonstrating DDE-based approach with weighted components of delayed arguments and instant transitions. Thus, we believe the proposed COVID-19 model and simulation results may be of interest to both experts in epidemiological modeling and a more general audience.

The final version of the DDE-based model still has a number of abstract parameters reflecting government NPIs, conducted testing procedure, imposing quarantine for those who were detected as infected with the virus and importing infected individuals into the modeling region or country. Particularly, parameters describing fractions of different symptoms’ severity should be correctly attributed to the patient’s age that requires extension of the model.

One of the crucial issues in the case of COVID-19 epidemiological modeling is to correctly transfer government NPIs (limit on mass gathering, lockdown, curfew, etc.) into the model parameters. One can easily see that introduction of a social distance” multiplier to the infection rate and fitting its value to experimental data enables it to reproduce almost any observable epidemiological trajectory. In order to tackle this problem we tried to utilize stringency index (Hale et al., 2020) instead of trying to fit the “social distance” factor over time of the pandemic in a certain country. However, we still have quite abstract aggregated numerical values of the indicator. Thus, further step for the model development in this direction is to use individual components or NPIs of the stringency index and attempt to assess their individual effect on the epidemic.

Additionally, we still do not entirely understand the reason behind the second and consequent waves. It is evident that the stringency index alone can not explain those waves in every country. In the model we tried to overcome these problems by means of changes in adherence to government NPIs and/or increase of the virus contagiousness. The second is explained by the emergence of new (more contagious) strains of the virus (Andersson et al., 2021; Jones et al., 2021; Kirby, 2021; Sheikh et al., 2021). However, the latter should be modeled more correctly using a modified version of the model with two or more similar modules where each of them is containing a full disease progressing scheme taking into account separate strains. This is also our plan in the development of the model. Moreover, the model fitting to statistical data for both countries demonstrated the decrease of the infection fatality rate during the COVID-19 epidemic which corresponds to early statements that the fatality rate of the Delta or B.1.617.2 variant of COVID-19, for example, is lower than the original variant. However, it might be caused by the age of unvaccinated people who were infected by the Delta virus strains and hospitalized with severe symptoms. According to the report published by Public Health England (Public Health England report, 2021), for instance, the majority of COVID-19 cases caused by the Delta variant were detected in people under 50 years old in the UK which are less likely to die from COVID-19 compared to those older than 50. So the current comparison of the case fatality rate of the B.1.617.2 variant with that of the wild-type virus is biased due to vaccination strategy in some European countries and age stratification of the population. This statistical misleading indicates the necessity to specify the developed model for each age group and integrate them in a more complex DDE model considering age distribution in a certain country. So the roadmap for the extension and further model development includes the next biological aspects of the virus and epidemiology of the COVID-19 pandemic:

- Age-specific modules (infection and death rates, hospitalization and severity of the disease, B and T cell response) according to the statistical data (Monod et al., 2020; Yang et al., 2021).
- Explicit strain emergence with other viral indicators like infectivity, resistance to neutralization, vaccine effectiveness in the population (Chen et al., 2021; Davies et al., 2021; Faria et al., 2021; Jalkanen et al., 2021; Sheikh et al., 2021; Volz et al., 2021; Washington et al., 2021; Zhou et al., 2021).
- Waning B and T cell immunity and neutralization activity of specific antibodies (Dan et al., 2021; Harrington et al., 2021; Sherina et al., 2021; Zuo et al., 2021).
- Effect of reinfection on the epidemic dynamics (Krutikov et al., 2021, Team C.C.-V.B.C.I., 2021).
- Superspreading events (Althouse et al., 2020; Lemieux et al., 2020; Yang et al., 2021a).

## Data Availability

The data that support the findings of this study are available from Our World In Data website. Datasets selected for particular regions are also available via the web interface of BioUML software accessible through dedicated gitlab page.

https://gitlab.sirius-web.org/covid-19/dde-epidemiology-model.

https://ourworldindata.org/

## Model availability

The developed model is available through the web interface of BioUML software (Kolpakov et al., 2020) at https://gitlab.sirius-web.org/covid-19/dde-epidemiology-model. Models are available both through visual representation in the platform and in Jupyter notebooks which allow users to reproduce simulation results and figures presented in this study.

## Acknowledgements

The study was supported financially by the RFBR (#20-04-60355 “Development of a multi-scale immuno-epidemiological mathematical model COVID-19, taking into account the impact on the economy of the region and the scenarios of authority actions”)

## References

1. Adam, D., 2020. Special report: The simulations driving the world’s response to COVID-19. Nature, 580(7802), pp.316–319.

2. Althouse, B.M., Wenger, E.A., Miller, J.C., Scarpino, S.V., Allard, A., Hébert-Dufresne, L. and Hu, H., 2020. Superspreading events in the transmission dynamics of SARS-CoV-2: Opportunities for interventions and control. PLoS biology, 18(11), p.e3000897.

3. Andersson, P., Sherry, N.L. and Howden, B.P., 2021. Surveillance for SARS-CoV-2 variants of concern in the Australian context. Medical Journal of Australia.

4. Arino J., Portet S. A simple model for COVID-19. Infectious Disease Modelling. 5. 2020. 309–315.

5. Atkeson, A., 2020. What will be the economic impact of COVID-19 in the US? Rough estimates of disease scenarios (No. w26867). National Bureau of Economic Research.

6. Banerjee, M., Tokarev, A. and Volpert, V., 2020. Immuno-epidemiological model of two-stage epidemic growth. Mathematical Modelling of Natural Phenomena, 15, p.27.

7. Boëlle, P.Y., Delory, T., Maynadier, X., Janssen, C., Piarroux, R., Pichenot, M., Lemaire, X., Baclet, N., Weyrich, P., Melliez, H. and Meybeck, A., 2020. Trajectories of hospitalization in COVID-19 patients: an observational study in France. Journal of clinical medicine, 9(10), p.3148.

8. Böhmer, M.M., Buchholz, U., Corman, V.M., Hoch, M., Katz, K., Marosevic, D.V., Böhm, S., Woudenberg, T., Ackermann, N., Konrad, R. and Eberle, U., 2020. Investigation of a COVID-19 outbreak in Germany resulting from a single travel-associated primary case: a case series. The Lancet Infectious Diseases, 20(8), pp.920–928.

9. Deslandes, A., Berti, V., Tandjaoui-Lambotte, Y., Alloui, C., Carbonnelle, E., Zahar, J.R., Brichler, S. and Cohen, Y., 2020. SARS-CoV-2 was already spreading in France in late December 2019. International journal of antimicrobial agents, 55(6), p.106006.

10. Calafiore, G.C., Novara, C. and Possieri, C., 2020. A time-varying SIRD model for the COVID-19 contagion in Italy. Annual reviews in control.

11. Carrat F., Figoni, J., Henny, J., Desenclos, J.C., Kab, S., de Lamballerie, X. and Zins, M., 2021. Evidence of early circulation of SARS-CoV-2 in France: findings from the population-based “CONSTANCES” cohort. European Journal of Epidemiology, 36(2), pp.219–222.

12. Chen, R.E., Zhang, X., Case, J.B., Winkler, E.S., Liu, Y., VanBlargan, L.A., Liu, J., Errico, J.M., Xie, X., Suryadevara, N. and Gilchuk, P., 2021. Resistance of SARS-CoV-2 variants to neutralization by monoclonal and serum-derived polyclonal antibodies. Nature Medicine, 27(4), pp.717–726.

13. Chinazzi, M., Davis, J.T., Ajelli, M., Gioannini, C., Litvinova, M., Merler, S., y Piontti, A.P., Mu, K., Rossi, L., Sun, K. and Viboud, C., 2020. The effect of travel restrictions on the spread of the 2019 novel coronavirus (COVID-19) outbreak. Science, 368(6489), pp.395–400.

14. Ciuriak, D. and Fay, R., 2020. The Critical Numbers Game: How Models can Inform the Pandemic Policy Response from Lockdown to Reboot. Opinion, Centre for International Governance Innovation.

15. Cobey, S., 2020. Modeling infectious disease dynamics. Science, 368(6492), pp.713–714.

16. Cooke, K.L. and Van Den Driessche, P., 1996. Analysis of an SEIRS epidemic model with two delays. Journal of Mathematical Biology, 35(2), pp.240–260.

17. Dan, J.M., Mateus, J., Kato, Y., Hastie, K.M., Yu, E.D., Faliti, C.E., Grifoni, A., Ramirez, S.I., Haupt, S., Frazier, A. and Nakao, C., 2021. Immunological memory to SARS-CoV-2 assessed for up to 8 months after infection. Science, 371(6529).

18. Davies, N.G., Jarvis, C.I., Edmunds, W.J., Jewell, N.P., Diaz-Ordaz, K. and Keogh, R.H., 2021. Increased mortality in community-tested cases of SARS-CoV-2 lineage B. 1.1. 7. Nature, 593(7858), pp.270–274.

19. Del Fava, E., Cimentada, J., Perrotta, D., Grow, A., Rampazzo, F., Gil-Clavel, S. and Zagheni, E., 2020. The differential impact of physical distancing strategies on social contacts relevant for the spread of COVID-19. medRxiv.

20. Devipriya R. Dhamodharavadhani S., Selvi S. SEIR Model for COVID-19 Epidemic Using Delay Differential Equation 2021 J. Phys.: Conf. Ser. 1767 012005.

21. Doshi, P., 2020. Covid-19: Do many people have pre-existing immunity? Bmj, 370.

22. European Commission. 2020a. “COVID-19: Temporary Restriction on Non-Essential Travel to the EU.” European Commission. accessed 16 March 2020a. https://eur-lex.europa.eu/legal-content/EN/TXT/HTML/?uri=CELEX:52020DC0115&from=EN.

23. Fanelli, D. and Piazza, F., 2020. Analysis and forecast of COVID-19 spreading in China, Italy and France. Chaos, Solitons & Fractals, 134, p.109761.

24. Faria, N.R., Mellan, T.A., Whittaker, C., Claro, I.M., Candido, D.D.S., Mishra, S., Crispim, M.A., Sales, F.C., Hawryluk, I., McCrone, J.T. and Hulswit, R.J., 2021. Genomics and epidemiology of the P. 1 SARS-CoV-2 lineage in Manaus, Brazil. Science, 372(6544), pp.815–821.

25. Ferguson, N.M., Laydon, D., Nedjati-Gilani, G., Imai, N., Ainslie, K., Baguelin, M., Bhatia, S., Boonyasiri, A., Cucunubá, Z., Cuomo-Dannenburg, G. and Dighe, A., 2020. Impact of non-pharmaceutical interventions (NPIs) to reduce COVID-19 mortality and healthcare demand. Imperial College COVID-19 Response Team. Imperial College COVID-19 Response Team, p.20.

26. Frampton, D., Rampling, T., Cross, A., Bailey, H., Heaney, J., Byott, M., Scott, R., Sconza, R., Price, J., Margaritis, M. and Bergstrom, M., 2021. Genomic characteristics and clinical effect of the emergent SARS-CoV-2 B. 1.1.7 lineage in London, UK: a whole-genome sequencing and hospital-based cohort study. The Lancet Infectious Diseases.

27. Götz, T. and Heidrich, P., 2020. Early stage COVID-19 disease dynamics in Germany: models and parameter identification. Journal of Mathematics in Industry, 10(1), pp.1–13.

28. Hale, T., Angrist, N., Goldszmidt, R., Kira, B., Petherick, A., Phillips, T., Webster, S., Cameron-Blake, E., Hallas, L., Majumdar, S. and Tatlow, H., 2021. A global panel database of pandemic policies (Oxford COVID-19 Government Response Tracker). Nature Human Behaviour, 5(4), pp.529–538.

29. Harrington, W.E., Trakhimets, O., Andrade, D.V., Dambrauskas, N., Raappana, A., Jiang, Y., Houck, J., Selman, W., Yang, A., Vigdorovich, V. and Yeung, W., 2021. Rapid decline of neutralizing antibodies is associated with decay of IgM in adults recovered from mild COVID-19. Cell Reports Medicine, 2(4), p.100253.

30. Hodcroft, E.B., 2021. CoVariants: SARS-CoV-2 Mutations and Variants of Interest. https://covariants.org/

31. Hoops, S., Sahle, S., Gauges, R., Lee, C., Pahle, J., Simus, N., Singhal, M., Xu, L., Mendes, P. and Kummer, U., 2006. COPASI—a complex pathway simulator. Bioinformatics, 22(24), pp.3067–3074.

32. Hucka, M., Bergmann, F.T., Dräger, A., Hoops, S., Keating, S.M., Le Novère, N., Myers, C.J., Olivier, B.G., Sahle, S., Schaff, J.C. and Smith, L.P., 2018. The Systems Biology Markup Language (SBML): language specification for level 3 version 2 core. Journal of integrative bioinformatics, 15(1).

33. Jalkanen, P., Kolehmainen, P., Häkkinen, H., Huttunen, M., Tähtinen, P., Lundberg, R., Maljanen, S., Reinholm, A., Tauriainen, S., Pakkanen, S. and Levonen, I., 2021. COVID-19 mRNA vaccine induced antibody responses and neutralizing antibodies against three SARS-CoV-2 variants. Nature Communications, 12, 3991.

34. Jones, T.C., Biele, G., Mühlemann, B., Veith, T., Schneider, J., Beheim-Schwarzbach, J., Bleicker, T., Tesch, J., Schmidt, M.L., Sander, L.E. and Kurth, F., 2021. Estimating infectiousness throughout SARS-CoV-2 infection course. Science. 373(6551), eabi5273.

35. Karagiannidis, C., Windisch, W., McAuley, D.F., Welte, T. and Busse, R., 2021. Major differences in ICU admissions during the first and second COVID-19 wave in Germany. The Lancet Respiratory Medicine, 9(5), pp.e47–e48..

36. Kermack, W.O. and McKendrick, A.G., 1927. A contribution to the mathematical theory of epidemics. Proceedings of the royal society of london. Series A, Containing papers of a mathematical and physical character, 115(772), pp.700–721.

37. Kirby, T., 2021. New variant of SARS-CoV-2 in UK causes surge of COVID-19. The Lancet Respiratory Medicine, 9(2), pp.e20–e21.

38. Kissler, S.M., Tedijanto, C., Goldstein, E., Grad, Y.H. and Lipsitch, M., 2020. Projecting the transmission dynamics of SARS-CoV-2 through the postpandemic period. Science, 368(6493), pp.860–868.

39. Kolpakov, F., Akberdin, I., Kashapov, T., Kiselev, L., Kolmykov, S., Kondrakhin, Y., Kutumova, E., Mandrik, N., Pintus, S., Ryabova, A. and Sharipov, R., 2019. BioUML: an integrated environment for systems biology and collaborative analysis of biomedical data. Nucleic acids research, 47(W1), pp.W225–W233.

40. Krutikov, M., Palmer, T., Tut, G., Fuller, C., Shrotri, M., Williams, H., Davies, D., Irwin-Singer, A., Robson, J., Hayward, A. and Moss, P., 2021. Incidence of SARS-CoV-2 infection according to baseline antibody status in staff and residents of 100 long-term care facilities (VIVALDI): a prospective cohort study. The Lancet Healthy Longevity, 2(6), pp.e362–e370.

41. Kucharski, A.J., Russell, T.W., Diamond, C., Liu, Y., Edmunds, J., Funk, S., Eggo, R.M., Sun, F., Jit, M., Munday, J.D. and Davies, N., 2020. Early dynamics of transmission and control of COVID-19: a mathematical modelling study. The Lancet Infectious Diseases, 20(5), pp.553–558.

42. Lauer SA, Grantz KH, Bi Q, et al. The Incubation Period of Coronavirus Disease 2019 (COVID-19) From Publicly Reported Confirmed Cases: Estimation and Application. Ann Intern Med. 2020;172(9):577–582.

43. Lemieux, J.E., Siddle, K.J., Shaw, B.M., Loreth, C., Schaffner, S.F., Gladden-Young, A., Adams, G., Fink, T., Tomkins-Tinch, C.H., Krasilnikova, L.A. and DeRuff, K.C., 2021. Phylogenetic analysis of SARS-CoV-2 in Boston highlights the impact of superspreading events. Science, 371(6529).

44. Le Novere, N., Hucka, M., Mi, H., Moodie, S., Schreiber, F., Sorokin, A., Demir, E., Wegner, K., Aladjem, M.I., Wimalaratne, S.M. and Bergman, F.T., 2009. The systems biology graphical notation. Nature biotechnology, 27(8), pp.735–741.

45. Li, M.Y. and Muldowney, J.S., 1995. Global stability for the SEIR model in epidemiology. Mathematical Biosciences, 125(2), pp.155–164.

46. Martcheva, M., 2015. An introduction to mathematical epidemiology (Vol. 61). New York: Springer.

47. Mateus, J., Grifoni, A., Tarke, A., Sidney, J., Ramirez, S.I., Dan, J.M., Burger, Z.C., Rawlings, S.A., Smith, D.M., Phillips, E. and Mallal, S., 2020. Selective and cross-reactive SARS-CoV-2 T cell epitopes in unexposed humans. Science, 370(6512), pp.89–94.

48. Menendez, J., 2020. Elementary time-delay dynamics of COVID-19 disease. medRxiv.

49. Metcalf, C.J.E., Morris, D.H. and Park, S.W., 2020. Mathematical models to guide pandemic response. Science, 369(6502), pp.368–369.

50. Monod, M., Blenkinsop, A., Xi, X., Hebert, D., Bershan, S., Tietze, S., Baguelin, M., Bradley, V.C., Chen, Y., Coupland, H. and Filippi, S., 2021. Age groups that sustain resurging COVID-19 epidemics in the United States. Science, 371(6536).

51. Ng, K.W., Faulkner, N., Cornish, G.H., Rosa, A., Harvey, R., Hussain, S., Ulferts, R., Earl, C., Wrobel, A.G., Benton, D.J. and Roustan, C., 2020. Preexisting and de novo humoral immunity to SARS-CoV-2 in humans. Science, 370(6522), pp.1339–1343.

52. Oran DP, Topol EJ. The Proportion of SARS-CoV-2 Infections That Are Asymptomatic : A Systematic Review. Ann Intern Med. 2021;174(5):655–662.

53. Pinto, D., Park, Y.J., Beltramello, M., Walls, A.C., Tortorici, M.A., Bianchi, S., Jaconi, S., Culap, K., Zatta, F., De Marco, A. and Peter, A., 2020. Cross-neutralization of SARS-CoV-2 by a human monoclonal SARS-CoV antibody. Nature, 583(7815), pp.290–295.

54. Prem, K., Liu, Y., Russell, T.W., Kucharski, A.J., Eggo, R.M., Davies, N., Flasche, S., Clifford, S., Pearson, C.A., Munday, J.D. and Abbott, S., 2020. The effect of control strategies to reduce social mixing on outcomes of the COVID-19 epidemic in Wuhan, China: a modelling study. The Lancet Public Health, 5(5), pp.e261–e270.

55. Public Health England, 2021. SARS-CoV-2 variants of concern and variants under investigation in England. Technical briefing 18. URL https://assets.publishing.service.gov.uk/government/uploads/system/uploads/attachment_data/file/1001358/Variants_of_Concern_VOC_Technical_Briefing_18.pdf. [Online] (Accessed: 22 July 2021).

56. Rimmelé, T., Pascal, L., Polazzi, S. and Duclos, A., 2021. Organizational aspects of care associated with mortality in critically ill COVID-19 patients. Intensive care medicine, 47(1), pp.119–121.

57. Sharma, N., Verma, A.K. and Gupta, A.K., 2021. Spatial network based model forecasting transmission and control of COVID-19. Physica A: Statistical Mechanics and its Applications, p.126223.

58. Shayak, B., Sharma, M.M., Rand, R.H., Singh, A. and Misra, A.N.O.O.P., 2020. A Delay differential equation model for the spread of COVID-19. International Journal of Engineering Research and Applications, 10(10/3), pp.1–13.

59. Sheikh, A., McMenamin, J., Taylor, B. and Robertson, C., 2021. SARS-CoV-2 Delta VOC in Scotland: demographics, risk of hospital admission, and vaccine effectiveness. The Lancet.

60. Sherina, N., Piralla, A., Du, L., Wan, H., Kumagai-Braesch, M., Andréll, J., Braesch-Andersen, S., Cassaniti, I., Percivalle, E., Sarasini, A. and Bergami, F., 2021. Persistence of SARS-CoV-2-specific B and T cell responses in convalescent COVID-19 patients 6–8 months after the infection. Med, 2(3), pp.281–295.

61. Shrock, E., Fujimura, E., Kula, T., Timms, R.T., Lee, I.H., Leng, Y., Robinson, M.L., Sie, B.M., Li, M.Z., Chen, Y. and Logue, J., 2020. Viral epitope profiling of COVID-19 patients reveals cross-reactivity and correlates of severity. Science, 370(6520).

62. Team, C.C.-V.B.C.I., 2021. COVID-19 Vaccine Breakthrough Infections Reported to CDC—United States, January 1–April 30, 2021. Morbidity and Mortality Weekly Report, 70(21), p.792.

63. Tuomisto, J.T., Yrjölä, J., Kolehmainen, M., Bonsdorff, J., Pekkanen, J. and Tikkanen, T., 2020. An agent-based epidemic model REINA for COVID-19 to identify destructive policies. medRxiv.

64. Utamura, M., Koizumi, M. and Kirikami, S., 2020. Isolation Considered Epidemiological Model for the Prediction of COVID-19 Trend in Tokyo, Japan: Numerical Study. JMIR public health and surveillance, 6(4).

65. Volz, E., Mishra, S., Chand, M., Barrett, J.C., Johnson, R., Geidelberg, L., Hinsley, W.R., Laydon, D.J., Dabrera, G., O’Toole, Á. and Amato, R., 2021. Assessing transmissibility of SARS-CoV-2 lineage B. 1.1. 7 in England. Nature, 593(7858), pp.266–269.

66. Washington, N.L., Gangavarapu, K., Zeller, M., Bolze, A., Cirulli, E.T., Barrett, K.M.S., Larsen, B.B., Anderson, C., White, S., Cassens, T. and Jacobs, S., 2021. Emergence and rapid transmission of SARS-CoV-2 B. 1.1. 7 in the United States. Cell, 184(10), pp.2587–2594.

67. Westerhoff, H.V. and Kolodkin, A.N., 2020. Advice from a systems-biology model of the corona epidemics. NPJ systems biology and applications, 6(1), pp.1–5.

68. WHO Situation Report 6 March. https://www.who.int/docs/default-source/coronaviruse/situation-reports/20200306-sitrep-46-covid-19.pdf?sfvrsn=96b04adf_4

69. Yang, Z., Zeng, Z., Wang, K., Wong, S.S., Liang, W., Zanin, M., Liu, P., Cao, X., Gao, Z., Mai, Z. and Liang, J., 2020. Modified SEIR and AI prediction of the epidemics trend of COVID-19 in China under public health interventions. Journal of thoracic disease, 12(3), p.165.

70. Yang, F., Nielsen, S.C., Hoh, R.A., Röltgen, K., Wirz, O.F., Haraguchi, E., Jean, G.H., Lee, J.Y., Pham, T.D., Jackson, K.J. and Roskin, K.M., 2021. Shared B cell memory to coronaviruses and other pathogens varies in human age groups and tissues. Science, 372(6543), pp.738–741.

71. Yang, Q., Saldi, T.K., Gonzales, P.K., Lasda, E., Decker, C.J., Tat, K.L., Fink, M.R., Hager, C.R., Davis, J.C., Ozeroff, C.D. and Muhlrad, D., 2021a. Just 2% of SARS-CoV-2− positive individuals carry 90% of the virus circulating in communities. Proceedings of the National Academy of Sciences, 118(21).

72. Zhang, Y., Su, X., Chen, W., Fei, C.N., Guo, L.R., Wu, X.L., Zhou, N., Guo, Y.T., Dong, X.C., Zhao, Y. and Wang, H.W., 2020. Epidemiological investigation on a cluster epidemic of COVID-19 in a collective workplace in Tianjin. Zhonghua liu xing bing xue za zhi= Zhonghua liuxingbingxue zazhi, 41(5), pp.649–653.

73. Zhou, D., Dejnirattisai, W., Supasa, P., Liu, C., Mentzer, A.J., Ginn, H.M., Zhao, Y., Duyvesteyn, H.M., Tuekprakhon, A., Nutalai, R. and Wang, B., 2021. Evidence of escape of SARS-CoV-2 variant B. 1.351 from natural and vaccine-induced sera. Cell, 184(9), pp.2348–2361.

74. Zuo, J., Dowell, A.C., Pearce, H., Verma, K., Long, H.M., Begum, J., Aiano, F., Amin-Chowdhury, Z., Hoschler, K., Brooks, T. and Taylor, S., 2021. Robust SARS-CoV-2-specific T cell immunity is maintained at 6 months following primary infection. Nature Immunology, 22(5), pp.620–626.

