## Supplementary materials for "A Delay Differential Equation approach to model the COVID-19 pandemic"

#### List of model equations

$$\frac{dS}{dt}=-TIC\cdot S-V(S,V_{day}), \qquad \frac{dE_O}{dt}=TIC\cdot S-K\cdot E_O+Import_{Rate},$$

$$\frac{dE}{dt}=(1-T_E)\cdot K\cdot E_O-F_1(E), \qquad \frac{dE^T}{dt}=T_E\cdot K\cdot E_O-F_1(E^T),$$

$$\frac{dA}{dt}=A_F\cdot F_1(E)-F_2(A), \qquad \frac{dA^T}{dt}=A_F\cdot F_1(E^T)-F_2(A^T),$$

$$\frac{dI_O}{dt}=(1-A_F)\cdot F_1(E)-K\cdot I_O, \qquad \frac{dI_O^T}{dt}=(1-A_F)\cdot F_1(E^T)+K\cdot T_I\cdot I_O-K\cdot I_O,^T$$

$$\frac{dI_R}{dt}=K\cdot(1-T_I)\cdot(1-H_F)\cdot M_O-F_3(I_R), \qquad \frac{dI_R^T}{dt}=K\cdot(1-T_I)\cdot(1-H_F)\cdot I_O^T-F_3(I_R^T),$$

$$\frac{dI_W}{dt}=K\cdot(1-T_I)\cdot H_F\cdot I_O-F_4(I_W), \qquad \frac{dI_W^T}{dt}=K\cdot(1-T_I)\cdot H_F\cdot I_O^T-F_4(I_W^T),$$

$$\frac{dH_O}{dt}=F_4(I_W)-K\cdot H_O, \qquad \frac{dH_O^T}{dt}=F_4(I_W^T)-K\cdot H_O^T+K\cdot T_H\cdot H_O,$$

$$\frac{dH_R}{dt}=K\cdot(1-T_H)\cdot(1-D_H)\cdot(1-C_F)\cdot H_O-F_5(H_R), \qquad \frac{dH_R^T}{dt}=K\cdot(1-D_H)\cdot(1-C_F)\cdot H_O^T-F_5(H_R^T),$$

$$\frac{dH_D}{dt}=K\cdot(1-T_H)\cdot D_H\cdot(1-C_F)H-F_6(H_D), \qquad \frac{dH_D^T}{dt}=K\cdot D_H\cdot(1-C_F)H^T-F_6(H_D^T),$$

$$\frac{dH_W}{dt}=K\cdot(1-T_H)\cdot(1-D_H)\cdot C_F\cdot H-F_7(H_W), \qquad \frac{dH_W^T}{dt}=K\cdot(1-D_H)\cdot C_F\cdot H^T-F_7(H_W^T),$$

$$\frac{dC}{dt}=F_7(H_W)+F_7(H_W^T)-C\cdot ICU\cdot ICU_{Rate}, \qquad \frac{dC_{ICU}}{dt}=C\cdot ICU\cdot ICU_{Rate}-F_8(C_{ICU}),$$

$$\frac{dR}{dt}=F_2(A)+F_3(I_R)+F_5(H_R), \qquad \frac{dR^T}{dt}=F_2(A^T)+F_3(I_R^T)+F_5(H_R^T)+(1-D_{ICU})\cdot F_8(C_{ICU}),$$

$$\frac{dD}{dt}=F_6(H_D)+F_8(C), \qquad \frac{dD^T}{dt}=F_6(H_D^T)+D_{ICU}\cdot F_8(C_{ICU}), \qquad \frac{dICU}{dt}=F_8(C_{ICU})-C\cdot ICU\cdot ICU_{Rate},$$

$$I_{total}=I_O+I_R+I_W, \quad H_{total}=H+H_R+H_W, \quad I_{total}^T=I^T+I_R^T+I_W^T, \quad H_{total}^T=H^T+H_R^T+H_W^T,$$

$$TIC=\frac{c\cdot(I_E\cdot(E+Q_E\cdot E^T)+I_A\cdot(A+A_T\cdot Q_A)+I_I\cdot(I_{total}+Q_I\cdot(I_{total}^T))+I_H\cdot(H_{total}+Q_H\cdot(H_{total}^T)))}{P}.$$

#### List of model variables and parameters

$S$ - susceptible subpopulation,
$P$ - total population,
$V$ - vaccinated,
$V_{day}$ - number of vaccinated per day,
$E_O$ - exposed, onset,
$E, E^T$ - exposed (not registered and registered as such),
$A, A^T$ - asymptomatic (not registered and registered),
$I_O, I_O^T$ - mild symptoms, onset (not registered and registered),
$I_R, I_R^T$ - mild symptoms, will recover (not registered and registered),
$I_W, I_W^T$ - mild symptoms will get worse (not registered and registered),
$I_{Total}, I_{Total}^T$ - mild symptoms, total (not registered and registered),
$H_O, H_O^T$ - severe symptoms, onset (not registered and registered),
$H_R, H_R^T$ - severe symptoms, will recover (not registered and registered),
$H_D, H_D^T$ - severe symptoms will die (not registered and registered),
$H_W, H_W^T$ - severe symptoms will transit to critical (not registered and registered),
$H_{Total}, H_{Total}^T$ - severe symptoms, total (not registered and registered),
$C$ - critical symptoms,
$C_{ICU}$ - critical symptoms, on ICU,
$ICU$ - number of free ICU,
$R, R^T$ - recovered (not registered and registered),
$D, D^T$ - deceased due to Covid-19 (not registered and registered),
$Import_{Rate}$ - number of infected arrived from other regions,
$TIC$ - Total Infection Coefficient,
$c$ - number of contacts per day,
$K$ - adjustment rate factor with significantly high value rendering reactions instant,
$I_A, I_E, I_I, I_H$ - infection rate of group,
$Q_A, Q_E, Q_I, Q_H$ - limiting of group upon testing,
$D_H, D_{ICU}$ - death rate in the group,
$T_E, T_I, T_H$ - testing in the group,
$A_F$ - fraction of asymptomatic among susceptible,
$H_F$ - fraction of severe symptoms among mild symptoms,
$CF$ - contacts level in the population,
$ICU_{Rate}$ - rate of transmission to the ICU.

### Fitted transition processes

Those charts as well as estimations may be reproduced using Jupyter notebook at https://sirius-web.org/bioumlweb/#de=data/Collaboration%20(git)/DDE%20Epidemiology%20model/Data/Jupyter/Processes%20estimation.ipynb

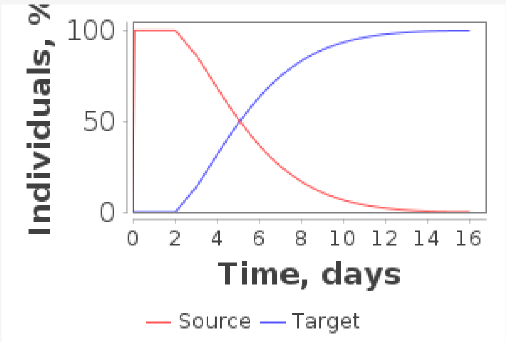


**Generated law:** 0.14*X( t - 2.0 ) + 0.04*X( t - 3.0 )

**Error:** 0.4041720918445345

**Quartiles:**

| Source | Q_25 | Q_1 | Q_2 | Q_3 | Q_975 |
|---|---|---|---|---|---|
| Model | 2.129 | 3.561 | 5.012 | 6.960 | 11.661 |
| Expected | 2.2 | 3.8 | 5.1 | 6.7 | 11.5 |

Supplementary Fig. 1. Incubation period process

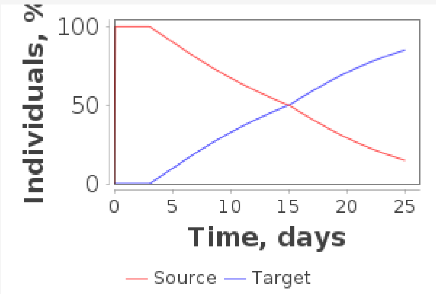


**Generated law:** 0.048*X( t - 3.0 ) + 0.0017*X( t - 4.0 ) + 0.0166*X( t - 15.0 )

**Error:** 1.5080651973149954

**Quartiles:**

| Source | Q_1 | Q_2 | Q_3 |
|---|---|---|---|
| Model | 8.127 | 14.936 | 21.176 |
| Expected | 8.0 | 14.0 | 20.0 |

Supplementary Fig.2. Recovery with mild or no symptoms

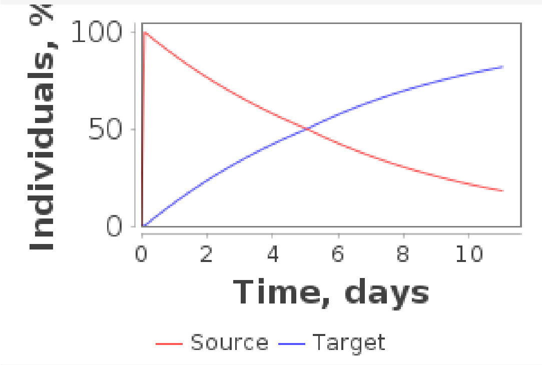


**Generated law:** 0.14*X( t - 0.0 ) + 0.014*X( t - 5.0 )

**Error:** 0.08068811198556733

**Quartiles:**

| Source | Q_1 | Q_2 | Q_3 |
|---|---|---|---|
| Model | 2.055 | 4.949 | 9.029 |
| Expected | 2.0 | 5.0 | 9.0 |

Supplementary Fig.3 Onset of severe symptoms

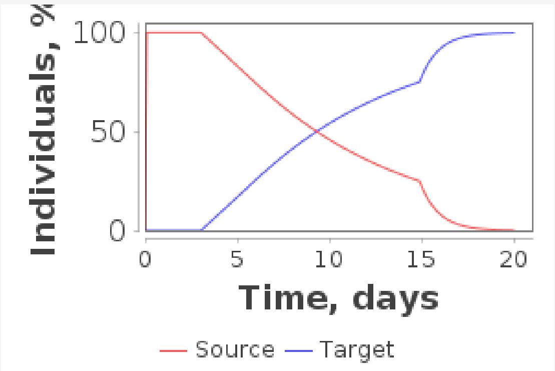


**Generated law:** 0.086*X( t - 3.0 ) + 0.67*X( t - 14.9 )

**Error:** 0.344036730176693

**Quartiles:**

| **Source** | **Q_1** | **Q_2** | **Q_3** |
|---|---|---|---|
| Model | 5.857 | 9.225 | 14.783 |
| Expected | 6.0 | 9.0 | 15.0 |

Supplementary Fig.4. Release from hospital without ICU (Boelle et al., 2020)

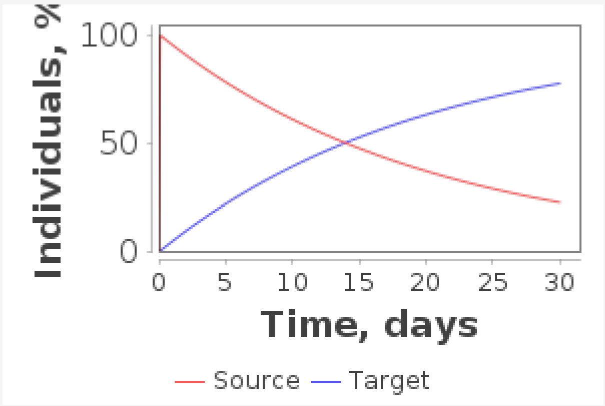


**Generated law:** 0.05*X( t - 0.0 )

**Error:** 0.0866556230775065З

**Quartiles:**

| **Source** | **Q_1** | **Q_2** | **Q_3** |
|---|---|---|---|
| Model | 5.754 | 13.863 | 27.726 |
| Expected | 5.7 | 13.8 | 27.7 |

Supplementary Fig.5. release from hospital without ICU (fitted on observed data in France)

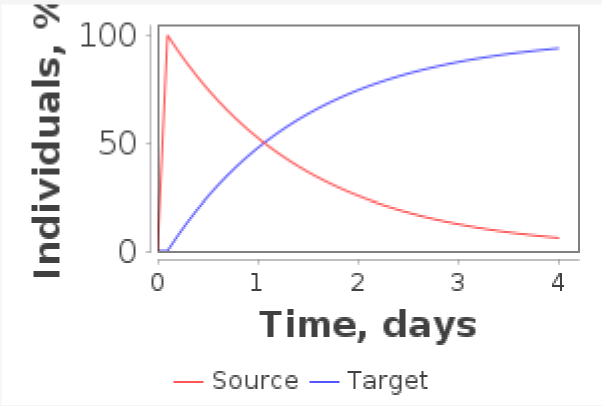


**Generated law:** 0.717*X( t - 0.0 )

**Error:** 0.40806782724803

**Quartiles:**

| **Source** | **Q_1** | **Q_2** | **Q_3** |
|---|---|---|---|
| Model | 0.401 | 0.967 | 1.933 |
| Expected | 0.0 | 1.0 | 2.0 |

Supplementary Fig.6. Onset of critical symptoms after severe

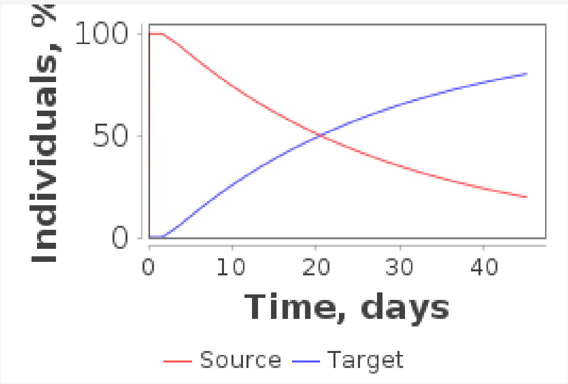


**Generated law:** 0.007*X( t - 3.583 ) + 0.028*X( t - 1.585 )

**Error:** 0.5749704266231722

**Quartiles:**

| Source | Q_1 | Q_2 | Q_3 |
|---|---|---|---|
| Model | 9.662 | 20.406 | 38.773 |
| Expected | 10.0 | 20.0 | 39.0 |

Supplementary Fig.7. Discharge from ICU
